# Determinants of malnutrition among Bangladeshi married women: A cross-sectional analysis of Bangladesh Demographic and Health Survey 2022

**DOI:** 10.1101/2025.08.24.25334328

**Authors:** Syeda Fahrima Shafrin, Mohammad Abu Tayeb Taki

**Affiliations:** Department of Economics, American International University-Bangladesh, Dhaka, Bangladesh; South Asia Gender Innovation Lab, World Bank, Dhaka, Bangladesh

## Abstract

**Background:** Malnutrition, encompassing both overnutrition and undernutrition, remains a global concern. In 2022, an estimated 2.5 billion adults worldwide were overweight and 390 million were underweight. In Bangladesh, women face a growing dual burden of malnutrition, with persistent undernutrition alongside rising overweight and obesity.

**Methods:** We conducted a cross-sectional analysis of data from 7,893 ever-married, non-pregnant women aged 15–49 years from the 2022 Bangladesh Demographic and Health Survey (BDHS). BMI was categorized as underweight (<18.5), normal (18.5–22.9), and overweight or obese (≥23) using WHO Asian guidelines. Survey-weighted multinomial logistic regression identified socio-demographic, reproductive, behavioral, and mental health factors associated with underweight and overweight or obesity, with normal weight as the reference.

**Results:** The weighted prevalence of normal weight, underweight and overweight or obesity was 33.7%, 9.5%, and 56.9%, respectively. Underweight was more common among adolescents, women from poorer and larger households, those experiencing depression, and those currently breastfeeding. Overweight or obesity was associated with older age, higher education, smaller and wealthier households, and sedentary behaviors such as frequent TV viewing and internet use. Later age at first birth and breastfeeding were protective against overweight or obesity. Depressed women were more likely to be underweight and less likely to be overweight or obese, while women with greater autonomy had higher odds of being overweight or obese.

**Conclusion:** Bangladeshi women face a pronounced dual burden of malnutrition, with overweight and obesity far exceeding underweight. Socio-demographic, behavioral, reproductive, and mental health factors play distinct roles, underscoring the need for multifaceted public health responses.

## Introduction

Malnutrition, encompassing both overnutrition and undernutrition, is a serious global concern. As per the World Bank reports, in 2022, approximately 2.5 billion adults worldwide were classified as overweight, while an estimated 390 million adults were underweight. This dual burden of malnutrition is not only a pressing issue in developing countries but also a matter of concern for developed nations.

While the socioeconomic patterns of malnutrition differ between high-income nations and low- and middle-income nations, the consequences are universally detrimental. In many low- and middle-income countries, obesity rates are highest in affluent urban populations [Dinsa, G. D., Goryakin, Y. – 2012], whereas in high-income countries lower-income groups bear the highest obesity burden [Hillier-Brown, F. C., Bambra, 2014]. Regardless of the contrasting patterns, both forms of malnutrition pose a serious threat to one’s physical and psychological health. In low- and middle-income countries, women are among the most vulnerable to malnutrition.

In Bangladesh, the nutritional landscape among women reflects a dual burden of malnutrition. According to [Khan S., Haider, 2022], 32% of ever-married women aged 15-49 were classified as overweight, while approximately 12% of women in the same age group were underweight. Overweight women are at an elevated risk for type 2 diabetes, hypertension, heart disease and hormone related cancers. [Pati, Irfan, Jameel, Ahmed, & Shahid, 2023]. Underweight women are more susceptible to infections, anemia, and face greater risk during childbirth due to low energy reserves. They face increased vulnerability to reproductive complications such as infertility, miscarriage, pre-term birth and low birth weight. [Sebire, Jolly, Harris, Regan, & Robinson, 2001]. While grappling with the dual burden of malnutrition, Bangladesh also faces major socioeconomic challenges like rising rates of child marriage and declining female labor force participation.

As per the UNICEF reports in 2023, the country ranks highest in South Asia for child marriage, with 51% of women aged 20 to 24 having been married before the age of 18. Early marriage often results in early pregnancies, compounding the health risks for already malnourished girls. This contributes to adverse birth outcomes such as low birth weight and stunting, initiating an intergenerational cycle of malnutrition. [Mim, S. A., Al Mamun, 2024]. Moreover, The HIES (Household and Income Expenditure Survey) 2022 reports that Bangladeshi women are less likely than men to participate in the labor force (42.5% vs. 81.3%). As a result, female labor force participation in Bangladesh is already quite low. The dual burden of malnutrition can further undermine women’s health, productivity, and ability to engage in the labor market. These socio-economic factors, combined with nutritional challenges, amplify the health and socio-economic vulnerabilities of women.

It is noteworthy that Bangladesh has made remarkable progress in reducing undernutrition rates in the past few years, yet the prevalence of overweight and obesity has been steadily increasing, creating a complex public health challenge [Hasan, Sutradhar, Shahabuddin, & Sarker, 2017]. Between 2007 to 2017–18, the prevalence of underweight among Bangladeshi women plummeted from 30% to 12%, while overweight and obesity jumped from 12% to 32% [Khan S., Haider, 2022]. As of 2018, only about one in eight women were underweight (down from nearly one in three in 2007), whereas one in three was above a healthy weight [Khan S., Haider, 2022]. These statistics illustrate that Bangladesh is experiencing a significant transition in the nutritional landscape among women. This transition requires a re-evaluation of public health strategies, urging the government to address the intricate interplay of factors contributing to both forms of malnutrition [Alem et al., 2023].

Several studies have investigated the underlying factors of malnutrition in ever-married women of age 15-49 in Bangladesh previously using Bangladesh Demographic and Health Survey (BDHS) data. Some of these studies have explored the determinants of either underweight or overweight separately [Rahman & Sathi, 2020], [Sarma et al., 2016], [Rahman, M. S., Mondal, 2015]. [Khan, M. M., & Kraemer, A. 2009], while many others have observed how the underlying factors of malnutrition differ in urban and rural settings. [Sumy et al., 2024], [Gupta, R. D., Frank, H. A., 2023]. Few more studies focus on examining both forms of malnutrition in women within a single analytical framework [Biswas, R. K., Rahman, N,2009], [Kamal, Hassan, & Alam, 2015], [Khanam, M., Osuagwu, 2021]. Despite the progress made, there is a lack of comprehensive evidence regarding the recent trends in malnutrition among Bangladeshi women based on the latest BDHS 2022 data. This study aims to bridge the gap by analyzing the emerging risk factors of malnutrition among married Bangladeshi women.

## Methods

### Data source and sampling procedure

Data were obtained from the latest round of the Bangladesh Demographic and Health survey [BDHS] 2022, which was the ninth nationally representative DHS survey conducted in Bangladesh. This study used de-identified secondary data from BDHS 2022, accessed in 27 June, 2025. No identifiable information was available to the authors.

BDHS is a nationally representative, cross-sectional survey conducted every five years to collect data on demographic, health, and nutritional indicators among women and children in Bangladesh. The 2022 BDHS was implemented through a collaborative effort of the National Institute of Population Research and Training (NIPORT) and ICF, with financial support from the Government of Bangladesh and USAID. This survey employed a two-stage stratified sampling design where in the first stage 675 enumeration areas [EAs] from each of Bangladesh’s eight administrative divisions (237 in urban areas and 438 in rural areas) were selected, with probability proportional to the unit size.

In the second stage, a systematic sample of 45 households was selected from each enumeration area. For the BDHS 2022 survey, four main questionnaires were used: The Household Questionnaire, the Woman’s Questionnaire (completed by ever-married women age 15–49), the Biomarker Questionnaire, and two Verbal Autopsy Questionnaires. This study is based upon the woman’s questionnaire which has a long and a short version. 30 of the 45 households in each EA were randomly selected for a long individual questionnaire and the rest of the 15 households were administered a short questionnaire.

Only 15 out of the 30 households with the long questionnaire were systematically selected for biomarker measurement, specifically height and weight measurements among ever-married women age 15-49.

A total of 30,330 households were surveyed out of which 30,078 women were interviewed. Afterwards, 10,053 women were selected for biomarker and full women questionnaire where height and weight related information were taken. The sample size stood at 9,100 ever-married women aged 15 to 49 years residing in the selected households, after excluding those who were pregnant or had given birth within the two months preceding the survey. Women with missing BMI values were dropped from the sample to ensure the accuracy of BMI calculations. [Sarma, H., Saquib, N., 2016], [Kamal, S. M., 2015], [Rahman, M. A., Rahman, 2019]. Finally, after dropping missing values in independent variables, the final sample size of this study became 7,893. Fig 1 given below describes the analytical sample selection process.

**Fig 1:**
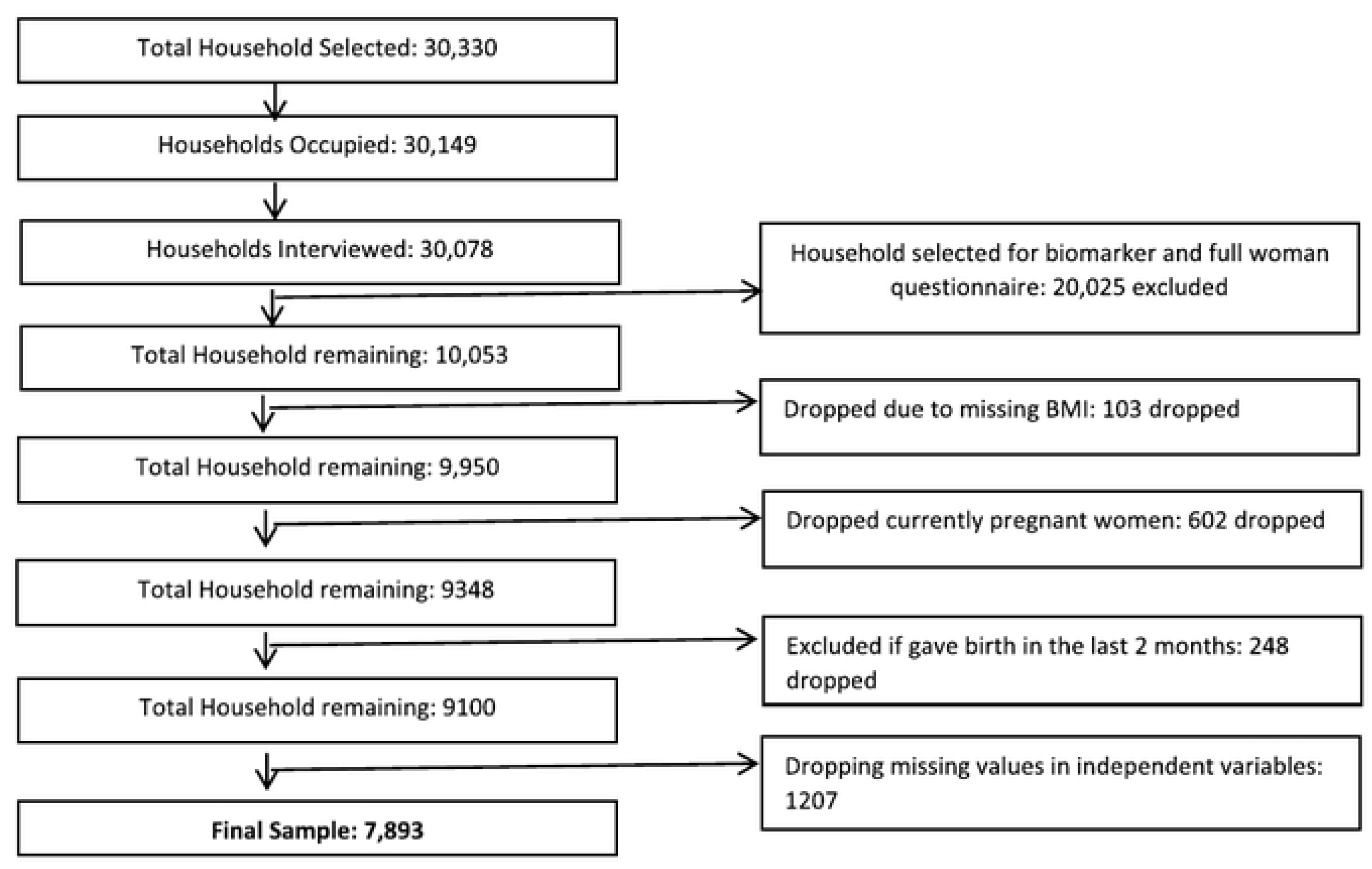
Diagram of Analytical Sample Selection Process

### Anthropometry measurements

A team of biomarker specialists were trained to ensure accuracy in measuring and recording height and weight data. Weight was measured using SECA digital scales (model number SECA 874U) and height was measured with a ShorrBoard® measuring board. Both instruments are standardized DHS equipments used globally for nutritional assessments.

### Ethical Consideration

Ethical approval for BDHS 2022 was obtained from the Institutional Review Board of ICF International, USA, and the Bangladesh Medical Research Council (BMRC). All participants provided written informed consent before participation; for respondents younger than 18 years, parental or guardian consent was obtained in addition to individual assent. The present study used secondary analysis of publicly available, de-identified data, and thus did not require separate ethical approval. The dataset is accessible from the DHS Program website. Date data were accessed for research on 27th June, 2025 Authors had no access to personally identifiable information. BDHS provides anonymized, publicly available datasets for research.

### Study variables

#### Outcome variable

The outcome variable was Body Mass Index [BMI] which was calculated as weight in kilograms divided by height in meters squared (kg/m²). BMI was categorized into three groups: underweight (BMI < 18.5), normal weight (18.5 ≤ BMI < 23), and overweight/obese (BMI ≥ 23) following the world Health Organization guidelines for Asian populations. [Tan KC, The Lancet 2004] Body mass index [BMI] was classified according to the Asian cut-off values, with normal weight as 18.5– 22.9 kg/m², underweight defined as BMI <18.5 kg/m² , and overweight or obese as ≥23 kg/m²; these categories were coded in Stata as 1, 2 and 3 respectively, with value labels assigned for analysis.

#### Explanatory variable

Drawing on existing research, theoretical considerations, and data availability/ Based on prior literature and data availability, the following covariates were considered: age group (15-19, 20-24, 25-29, 30-34, 35-39, 40-44,45-49), highest educational status (no formal education, primary, secondary, higher), wealth index (poorest, poorer, middle, richer, richest), parity (no children, 1-2, 3-4, 5+), place of residence (urban, rural), household size (small [1-3] , medium [4-6], large [7+] ]) currently in union/living with a man (yes, no), depression level (no depression, depression), anxiety level (no anxiety, anxiety), use of internet (not at all, yes-last 12 months, yes-before last 12 months), watching TV (not at all, at least once a week), currently breastfeeding [yes, no], birth in past one year,(no birth, at least 1 birth), age at first birth ( >20 years, <20 years ), pregnancy losses (no loss, at least one loss), use of contraceptive pills (yes, no), women’s agency (categorized as low, mild, full).

To calculate the wealth index, scores were assigned to each household based on its assets such as television, bicycles and cars; housing materials; and access to water and sanitation facilities. These scores were derived using principal component analysis (PCA). Subsequently, each person living in the household was assigned a score and based on this score, they were ranked into one of the five wealth quintiles.

BDHS 2022 included a mental health module for the first time to assess the level of depression and anxiety among ever married women age 15-49. Generalized Anxiety Disorder 7 scale (GAD-7) was used to evaluate symptoms of anxiety which classified anxiety severity based on the range of score 0 to 21. The scores were further classified as mild (0–5), moderate (6–14), and severe (15–21) [Spitzer, Kroenke, Williams, & Löwe, 2006] Patient Health Questionnaire, or PHQ-9 score was used to assess the symptoms of depression. Based on the range of PHQ score from 0 to 27, depression was categorized as minimal (0–4), mild (5–9), moderate (10–14), moderately severe (15–19), and severe (20–27). [Kroenke, Spitzer, & Williams, 2001] For the purpose of this study, scores of 6 or higher on the GAD-7 were categorized as “Anxiety,” and scores of 10 or higher on the PHQ-9 were categorized as “Depression”.To construct a robust index of women’s decision-making agency, four binary indicators were selected from the BDHS 2022 dataset: decision-making power over the respondent’s own health care, large household purchases, visits to family or relatives, and control over the husband’s earnings. Each variable was coded as 1 if the respondent had sole or joint decision-making power, and 0 otherwise. A Principal Component Analysis (PCA) was initially performed to derive a composite agency index, retaining the first component that explained the largest share of variance for use as a continuous score. However, due to limited variation in the PCA scores, we instead constructed a simple additive index, categorizing agency as 1 = Low, 2–3 = Mild, and 4 = Full agency. This approach is widely adopted in DHS-based studies.

### Statistical analysis

Descriptive analyses were initially performed to generate frequencies and percentages for all study variables Table 1. Given that the Bangladesh DHS follows a stratified, clustered survey design, sampling weights were applied to account for unequal probabilities of selection, clustering, and stratification.

**Table 1.**
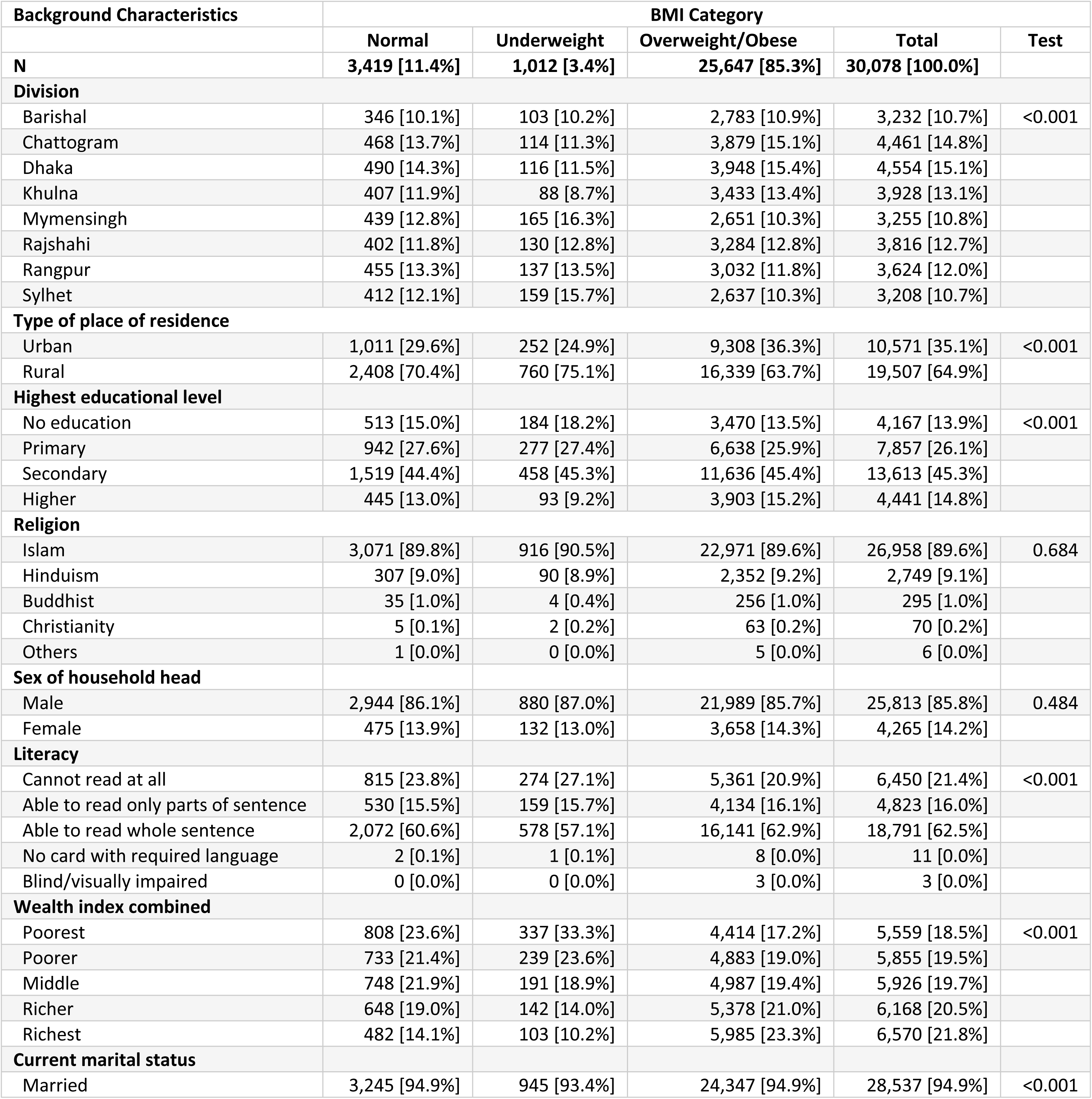
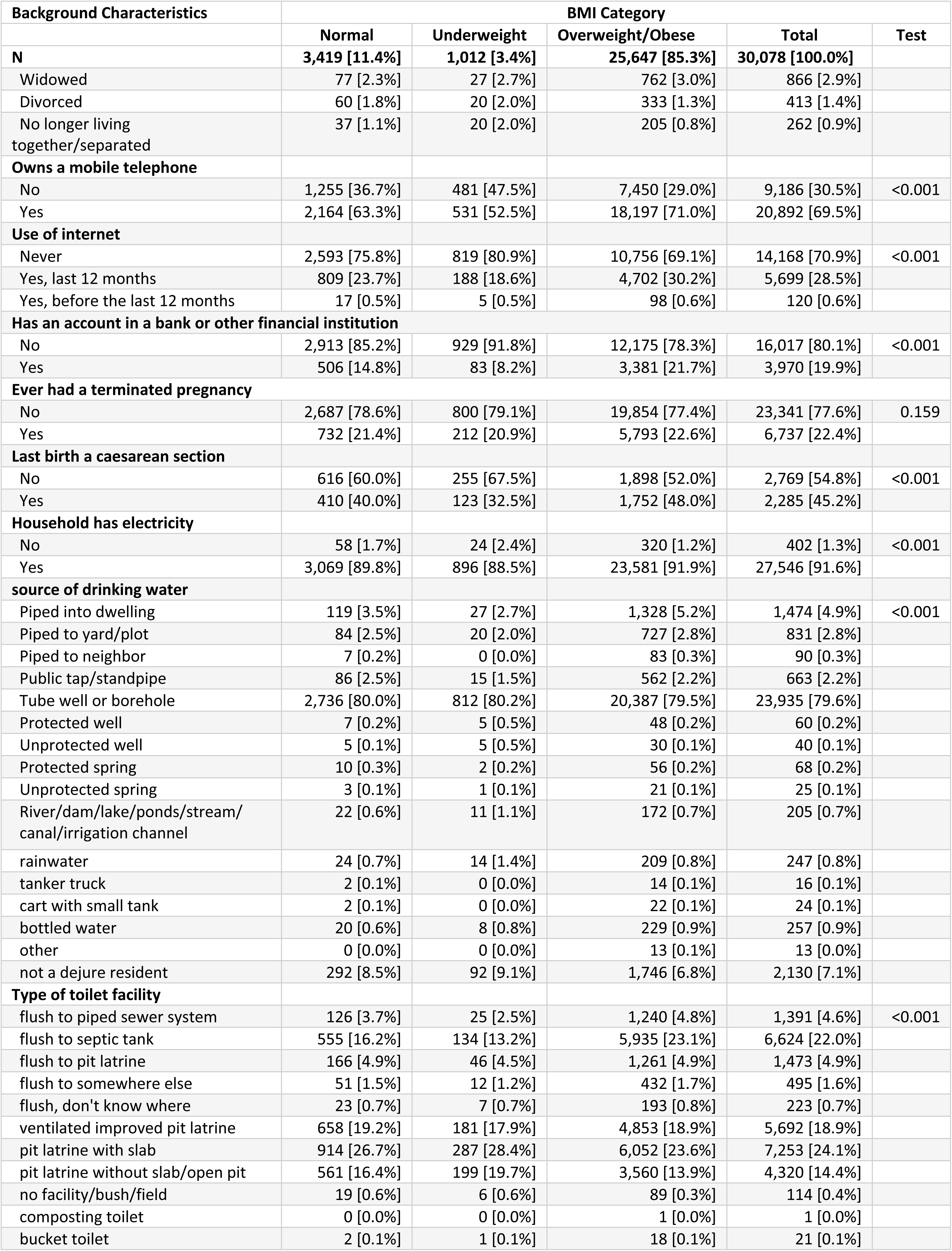
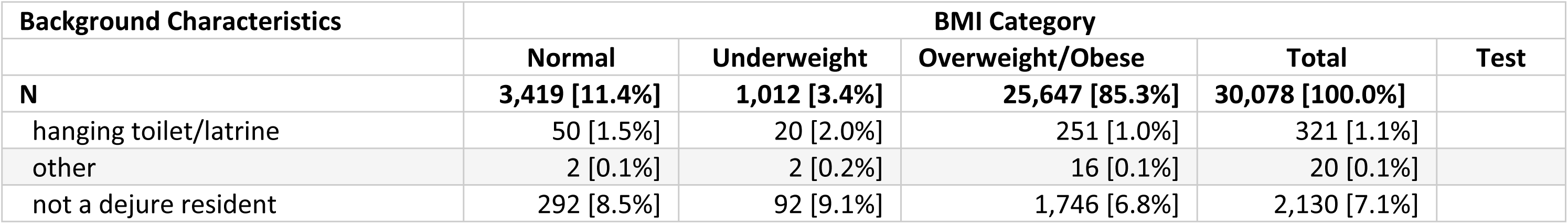
Distribution of married women across categories of BMI by different predictors, BDHS 2022.

| Background Characteristics | BMI Category |  |  |  |  |
| --- | --- | --- | --- | --- | --- |
|  | Normal | Underweight | Overweight/Obese | Total | Test |
| N | 3,419 [11.4%] | 1,012 [3.4%] | 25,647 [85.3%] | 30,078 [100.0%] |  |
| Division |  |  |  |  |  |
| Barishal | 346 [10.1%] | 103 [10.2%] | 2,783 [10.9%] | 3,232 [10.7%] | <0.001 |
| Chattogram | 468 [13.7%] | 114 [11.3%] | 3,879 [15.1%] | 4,461 [14.8%] |  |
| Dhaka | 490 [14.3%] | 116 [11.5%] | 3,948 [15.4%] | 4,554 [15.1%] |  |
| Khulna | 407 [11.9%] | 88 [8.7%] | 3,433 [13.4%] | 3,928 [13.1%] |  |
| Mymensingh | 439 [12.8%] | 165 [16.3%] | 2,651 [10.3%] | 3,255 [10.8%] |  |
| Rajshahi | 402 [11.8%] | 130 [12.8%] | 3,284 [12.8%] | 3,816 [12.7%] |  |
| Rangpur | 455 [13.3%] | 137 [13.5%] | 3,032 [11.8%] | 3,624 [12.0%] |  |
| Sylhet | 412 [12.1%] | 159 [15.7%] | 2,637 [10.3%] | 3,208 [10.7%] |  |
| Type of place of residence |  |  |  |  |  |
| Urban | 1,011 [29.6%] | 252 [24.9%] | 9,308 [36.3%] | 10,571 [35.1%] | <0.001 |
| Rural | 2,408 [70.4%] | 760 [75.1%] | 16,339 [63.7%] | 19,507 [64.9%] |  |
| Highest educational level |  |  |  |  |  |
| No education | 513 [15.0%] | 184 [18.2%] | 3,470 [13.5%] | 4,167 [13.9%] | <0.001 |
| Primary | 942 [27.6%] | 277 [27.4%] | 6,638 [25.9%] | 7,857 [26.1%] |  |
| Secondary | 1,519 [44.4%] | 458 [45.3%] | 11,636 [45.4%] | 13,613 [45.3%] |  |
| Higher | 445 [13.0%] | 93 [9.2%] | 3,903 [15.2%] | 4,441 [14.8%] |  |
| Religion |  |  |  |  |  |
| Islam | 3,071 [89.8%] | 916 [90.5%] | 22,971 [89.6%] | 26,958 [89.6%] | 0.684 |
| Hinduism | 307 [9.0%] | 90 [8.9%] | 2,352 [9.2%] | 2,749 [9.1%] |  |
| Buddhist | 35 [1.0%] | 4 [0.4%] | 256 [1.0%] | 295 [1.0%] |  |
| Christianity | 5 [0.1%] | 2 [0.2%] | 63 [0.2%] | 70 [0.2%] |  |
| Others | 1 [0.0%] | 0 [0.0%] | 5 [0.0%] | 6 [0.0%] |  |
| Sex of household head |  |  |  |  |  |
| Male | 2,944 [86.1%] | 880 [87.0%] | 21,989 [85.7%] | 25,813 [85.8%] | 0.484 |
| Female | 475 [13.9%] | 132 [13.0%] | 3,658 [14.3%] | 4,265 [14.2%] |  |
| Literacy |  |  |  |  |  |
| Cannot read at all | 815 [23.8%] | 274 [27.1%] | 5,361 [20.9%] | 6,450 [21.4%] | <0.001 |
| Able to read only parts of sentence | 530 [15.5%] | 159 [15.7%] | 4,134 [16.1%] | 4,823 [16.0%] |  |
| Able to read whole sentence | 2,072 [60.6%] | 578 [57.1%] | 16,141 [62.9%] | 18,791 [62.5%] |  |
| No card with required language | 2 [0.1%] | 1 [0.1%] | 8 [0.0%] | 11 [0.0%] |  |
| Blind/visually impaired | 0 [0.0%] | 0 [0.0%] | 3 [0.0%] | 3 [0.0%] |  |
| Wealth index combined |  |  |  |  |  |
| Poorest | 808 [23.6%] | 337 [33.3%] | 4,414 [17.2%] | 5,559 [18.5%] | <0.001 |
| Poorer | 733 [21.4%] | 239 [23.6%] | 4,883 [19.0%] | 5,855 [19.5%] |  |
| Middle | 748 [21.9%] | 191 [18.9%] | 4,987 [19.4%] | 5,926 [19.7%] |  |
| Richer | 648 [19.0%] | 142 [14.0%] | 5,378 [21.0%] | 6,168 [20.5%] |  |
| Richest | 482 [14.1%] | 103 [10.2%] | 5,985 [23.3%] | 6,570 [21.8%] |  |
| Current marital status |  |  |  |  |  |
| Married | 3,245 [94.9%] | 945 [93.4%] | 24,347 [94.9%] | 28,537 [94.9%] | <0.001 |

| Background Characteristics | BMI Category |  |  |  | Test |
| --- | --- | --- | --- | --- | --- |
|  | Normal | Underweight | Overweight/Obese | Total |  |
| **N** | **3,419 [11.4%]** | **1,012 [3.4%]** | **25,647 [85.3%]** | **30,078 [100.0%]** |  |
| Widowed | 77 [2.3%] | 27 [2.7%] | 762 [3.0%] | 866 [2.9%] |  |
| Divorced | 60 [1.8%] | 20 [2.0%] | 333 [1.3%] | 413 [1.4%] |  |
| No longer living together/separated | 37 [1.1%] | 20 [2.0%] | 205 [0.8%] | 262 [0.9%] |  |
| **Owns a mobile telephone** |  |  |  |  |  |
| No | 1,255 [36.7%] | 481 [47.5%] | 7,450 [29.0%] | 9,186 [30.5%] | <0.001 |
| Yes | 2,164 [63.3%] | 531 [52.5%] | 18,197 [71.0%] | 20,892 [69.5%] |  |
| **Use of internet** |  |  |  |  |  |
| Never | 2,593 [75.8%] | 819 [80.9%] | 10,756 [69.1%] | 14,168 [70.9%] | <0.001 |
| Yes, last 12 months | 809 [23.7%] | 188 [18.6%] | 4,702 [30.2%] | 5,699 [28.5%] |  |
| Yes, before the last 12 months | 17 [0.5%] | 5 [0.5%] | 98 [0.6%] | 120 [0.6%] |  |
| **Has an account in a bank or other financial institution** |  |  |  |  |  |
| No | 2,913 [85.2%] | 929 [91.8%] | 12,175 [78.3%] | 16,017 [80.1%] | <0.001 |
| Yes | 506 [14.8%] | 83 [8.2%] | 3,381 [21.7%] | 3,970 [19.9%] |  |
| **Ever had a terminated pregnancy** |  |  |  |  |  |
| No | 2,687 [78.6%] | 800 [79.1%] | 19,854 [77.4%] | 23,341 [77.6%] | 0.159 |
| Yes | 732 [21.4%] | 212 [20.9%] | 5,793 [22.6%] | 6,737 [22.4%] |  |
| **Last birth a caesarean section** |  |  |  |  |  |
| No | 616 [60.0%] | 255 [67.5%] | 1,898 [52.0%] | 2,769 [54.8%] | <0.001 |
| Yes | 410 [40.0%] | 123 [32.5%] | 1,752 [48.0%] | 2,285 [45.2%] |  |
| **Household has electricity** |  |  |  |  |  |
| No | 58 [1.7%] | 24 [2.4%] | 320 [1.2%] | 402 [1.3%] | <0.001 |
| Yes | 3,069 [89.8%] | 896 [88.5%] | 23,581 [91.9%] | 27,546 [91.6%] |  |
| **source of drinking water** |  |  |  |  |  |
| Piped into dwelling | 119 [3.5%] | 27 [2.7%] | 1,328 [5.2%] | 1,474 [4.9%] | <0.001 |
| Piped to yard/plot | 84 [2.5%] | 20 [2.0%] | 727 [2.8%] | 831 [2.8%] |  |
| Piped to neighbor | 7 [0.2%] | 0 [0.0%] | 83 [0.3%] | 90 [0.3%] |  |
| Public tap/standpipe | 86 [2.5%] | 15 [1.5%] | 562 [2.2%] | 663 [2.2%] |  |
| Tube well or borehole | 2,736 [80.0%] | 812 [80.2%] | 20,387 [79.5%] | 23,935 [79.6%] |  |
| Protected well | 7 [0.2%] | 5 [0.5%] | 48 [0.2%] | 60 [0.2%] |  |
| Unprotected well | 5 [0.1%] | 5 [0.5%] | 30 [0.1%] | 40 [0.1%] |  |
| Protected spring | 10 [0.3%] | 2 [0.2%] | 56 [0.2%] | 68 [0.2%] |  |
| Unprotected spring | 3 [0.1%] | 1 [0.1%] | 21 [0.1%] | 25 [0.1%] |  |
| River/dam/lake/ponds/stream/canal/irrigation channel | 22 [0.6%] | 11 [1.1%] | 172 [0.7%] | 205 [0.7%] |  |
| rainwater | 24 [0.7%] | 14 [1.4%] | 209 [0.8%] | 247 [0.8%] |  |
| tanker truck | 2 [0.1%] | 0 [0.0%] | 14 [0.1%] | 16 [0.1%] |  |
| cart with small tank | 2 [0.1%] | 0 [0.0%] | 22 [0.1%] | 24 [0.1%] |  |
| bottled water | 20 [0.6%] | 8 [0.8%] | 229 [0.9%] | 257 [0.9%] |  |
| other | 0 [0.0%] | 0 [0.0%] | 13 [0.1%] | 13 [0.0%] |  |
| not a de jure resident | 292 [8.5%] | 92 [9.1%] | 1,746 [6.8%] | 2,130 [7.1%] |  |
| **Type of toilet facility** |  |  |  |  |  |
| flush to piped sewer system | 126 [3.7%] | 25 [2.5%] | 1,240 [4.8%] | 1,391 [4.6%] | <0.001 |
| flush to septic tank | 555 [16.2%] | 134 [13.2%] | 5,935 [23.1%] | 6,624 [22.0%] |  |
| flush to pit latrine | 166 [4.9%] | 46 [4.5%] | 1,261 [4.9%] | 1,473 [4.9%] |  |
| flush to somewhere else | 51 [1.5%] | 12 [1.2%] | 432 [1.7%] | 495 [1.6%] |  |
| flush, don't know where | 23 [0.7%] | 7 [0.7%] | 193 [0.8%] | 223 [0.7%] |  |
| ventilated improved pit latrine | 658 [19.2%] | 181 [17.9%] | 4,853 [18.9%] | 5,692 [18.9%] |  |
| pit latrine with slab | 914 [26.7%] | 287 [28.4%] | 6,052 [23.6%] | 7,253 [24.1%] |  |
| pit latrine without slab/open pit | 561 [16.4%] | 199 [19.7%] | 3,560 [13.9%] | 4,320 [14.4%] |  |
| no facility/bush/field | 19 [0.6%] | 6 [0.6%] | 89 [0.3%] | 114 [0.4%] |  |
| composting toilet | 0 [0.0%] | 0 [0.0%] | 1 [0.0%] | 1 [0.0%] |  |
| bucket toilet | 2 [0.1%] | 1 [0.1%] | 18 [0.1%] | 21 [0.1%] |  |
|  | Normal | Underweight | Overweight/Obese | Total | Test |
| **N** | **3,419 [11.4%]** | **1,012 [3.4%]** | **25,647 [85.3%]** | **30,078 [100.0%]** |  |
| hanging toilet/latrine | 50 [1.5%] | 20 [2.0%] | 251 [1.0%] | 321 [1.1%] |  |
| other | 2 [0.1%] | 2 [0.2%] | 16 [0.1%] | 20 [0.1%] |  |
| not a de jure resident | 292 [8.5%] | 92 [9.1%] | 1,746 [6.8%] | 2,130 [7.1%] |  |

Weighted descriptive statistics were thus produced to ensure nationally representative estimates.

When building a regression model, there’s always a balance to consider. Including more variables can help explain the outcome better but adding too many can make the estimates less stable. The best model is the one that captures the most important relationships without becoming unnecessarily complicated. Following this, bivariate analyses were conducted using the chi-square test to assess associations between the independent variables and BMI categories. In addition, unadjusted bivariate logistic regression models were estimated to examine potential factors associated with the outcome variable [BMI categories]. Variables with a p-value of <0.20 in the bivariate analyses were selected for inclusion in the multivariable model, a threshold chosen to reduce residual confounding [Maldonado & Greenland, 1993]. The study categorized the dependent variable, Bmi_cat’, into 3 mutually exclusive groups which carry different implications in public health such as normal weight, underweight and overweight or obese. Because the sample included only a small proportion of obese women, the obese and overweight categories were combined. The study performed a survey-weight adjusted multivariable multinomial logistic regression treating normal BMI as the base or reference category.

Two unadjusted models were compared for model fitness using Akaike Information Criterion (AIC), Bayesian Information Criterion [BIC], and Adjusted Akaike Information Criterion (AAIC). The model with the lowest values of these criteria—indicating better goodness of fit—was retained for the final analysis. The odds ratio and goodness of fit were conducted in the final model without survey weights, but the final Multinomial regression model was survey weight adjusted to estimate the coefficients and the marginal effects influencing malnutrition in married non-pregnant women.

Multicollinearity among covariates was assessed using the Variance Inflation Factor (VIF). No significant multicollinearity was detected. The mean VIF for all variables was below 2.75.

A total of 7,893 women were interviewed, representing approximately 7,889 women in the target population after applying survey weights. This small difference reflects the adjustment made by the weights to ensure representativeness and is a normal feature of weighted survey data. The final model, incorporating a range of factors influencing women’s nutritional status, was evaluated using both marginal effects and relative risk ratios [RRRs] with corresponding 95% confidence intervals [CIs] to quantify the associations between explanatory variables and BMI categories. Statistical significance was set at the 5% level [p < 0.05]. The statistical analyses were conducted using Stata version 18.0 StataCorp, USA with ‘svy’ command to adjust for sampling weights, clustering effects and stratification.

## Findings

### Characteristics of the population under study

In the BDHS 2022 survey, 30,078 households were included in this nationally representative sample. The population was predominantly Muslim [89.6%], with 95% of respondents being married women. Just over one-third (35.1%) of participants resided in urban areas. Educational attainment remained relatively low, with 21.4% of women reporting no formal education and only 14.8% having completed education beyond the secondary level. Access to digital services was limited: 30.5% of women did not own a mobile phone, 70.9% had never used the internet, and 80.1% did not have an account in a bank or other financial institution. In contrast, access to basic utilities was comparatively high, with 79.5% of households using tube wells as their primary water source and only 1.3% reported no access to electricity—figures consistent with those reported in international literature. To examine how these characteristics relate to women’s nutritional status, we first conducted bivariate analyses using chi-square tests to assess the association between BMI category and each explanatory variable. These results have been illustrated in Table 1.

### Descriptive Statistics depicting the prevalence of the determinants in our sample

In our sample (n=7893), the prevalence of underweight, normal and overweight has been found to be 9.5%, 33,6% and 56.9% respectively. Overweight BMI is the highest around 42% in women aged between 30-39 years. The double burden is also quite evident in our empirical analysis, as we can see that the prevalence of being overweight and obese increases as we move to richer households.

Adversely, as we go down the income ladder that is towards poorer households the proportion of underweight BMI increases. Around 35% of women are living in urban areas, 66% of women living in urban areas are overweight and obese but only 6% of them are underweight. These results have been illustrated in Table 2. Interestingly, the prevalence of depression of women in the sample was around 5%. More than half of these depressed women were found to be overweight and obese, which is way higher than the people of normal BMI. The prevalence of women with autonomy of decision-making, such as taking important decisions related to the respondent’s health, large asset purchases of the household, visit to the relatives and the husband’s earnings are either taken solely by the respondent or jointly with partner, revealed that the prevalence of women’s agency or decision-making autonomy is 54.4%.

**Table 2.** Summary statistics illustrating the prevalence of determinants by study factors of married women aged 15-49 Bangladesh.

|  | BMI Category |  |  |  |  |
| --- | --- | --- | --- | --- | --- |
|  | Normal | Underweight | Overweight_Obese | Total | P-value |
| N | 2,655 [33.6%] | 747 [9.5%] | 4,487 [56.9%] | 7,889 [100.0%] |  |
| **Age in 5-year groups** |  |  |  |  |  |
| 15-19 | 163 [6.2%] | 84 [11.3%] | 71 [1.6%] | 318 [4.0%] | <0.001 |
| 20-24 | 506 [19.0%] | 166 [22.2%] | 440 [9.8%] | 1,112 [14.1%] |  |
| 25-29 | 465 [17.5%] | 135 [18.1%] | 762 [17.0%] | 1,362 [17.3%] |  |
| 30-34 | 472 [17.8%] | 103 [13.8%] | 921 [20.5%] | 1,497 [19.0%] |  |
| 35-39 | 447 [16.9%] | 97 [13.0%] | 969 [21.6%] | 1,514 [19.2%] |  |
| 40-44 | 334 [12.6%] | 78 [10.4%] | 734 [16.4%] | 1,146 [14.5%] |  |
| 45-49 | 267 [10.1%] | 84 [11.2%] | 590 [13.1%] | 941 [11.9%] |  |
| **Highest educational level** |  |  |  |  |  |
| No education | 426 [16.1%] | 140 [18.7%] | 568 [12.7%] | 1,133 [14.4%] | <0.001 |
| Primary | 770 [29.0%] | 225 [30.2%] | 1,185 [26.4%] | 2,181 [27.6%] |  |
| Secondary | 1,181 [44.5%] | 325 [43.4%] | 2,106 [46.9%] | 3,611 [45.8%] |  |
| Higher | 277 [10.4%] | 58 [7.7%] | 629 [14.0%] | 964 [12.2%] |  |
| **Wealth index combined** |  |  |  |  |  |
| Poorest | 639 [24.1%] | 242 [32.4%] | 548 [12.2%] | 1,429 [18.1%] | <0.001 |
| Poorer | 561 [21.1%] | 198 [26.5%] | 798 [17.8%] | 1,557 [19.7%] |  |
| Middle | 613 [23.1%] | 137 [18.3%] | 905 [20.2%] | 1,655 [21.0%] |  |
| Richer | 495 [18.6%] | 100 [13.4%] | 1,047 [23.3%] | 1,641 [20.8%] |  |
| Richest | 347 [13.1%] | 70 [9.4%] | 1,190 [26.5%] | 1,607 [20.4%] |  |
| **Type of place of residence** |  |  |  |  |  |
| Urban | 630 [23.7%] | 132 [17.7%] | 1,453 [32.4%] | 2,216 [28.1%] | <0.001 |
| Rural | 2,024 [76.3%] | 615 [82.3%] | 3,034 [67.6%] | 5,674 [71.9%] |  |
| **Household size** |  |  |  |  |  |
| Small [1–3] | 544 [20.5%] | 133 [17.8%] | 1,048 [23.4%] | 1,726 [21.9%] | <0.001 |
| Medium [4–6] | 1,611 [60.7%] | 443 [59.3%] | 2,720 [60.6%] | 4,774 [60.5%] |  |
| Large [7+] | 499 [18.8%] | 171 [22.9%] | 719 [16.0%] | 1,390 [17.6%] |  |
| **Depression level** |  |  |  |  |  |
| No Depression | 2,525 [95.1%] | 698 [93.4%] | 4,273 [95.2%] | 7,495 [95.0%] | 0.146 |
| Depression | 130 [4.9%] | 50 [6.6%] | 215 [4.8%] | 394 [5.0%] |  |
| **Use of internet** |  |  |  |  |  |
| Never | 2,049 [77.2%] | 622 [83.2%] | 3,095 [69.0%] | 5,766 [73.1%] | <0.001 |
| Yes, last 12 months | 594 [22.4%] | 119 [16.0%] | 1,368 [30.5%] | 2,082 [26.4%] |  |
| Yes, before last 12 months | 11 [0.4%] | 6 [0.8%] | 25 [0.5%] | 42 [0.5%] |  |
| **TV watching frequency** |  |  |  |  |  |
| Not at all | 1,321 [49.8%] | 396 [53.0%] | 1,765 [39.3%] | 3,482 [44.1%] | <0.001 |
| At least once a week or less | 1,334 [50.2%] | 352 [47.0%] | 2,722 [60.7%] | 4,408 [55.9%] |  |
| **Currently breastfeeding** |  |  |  |  |  |
| No | 1,881 [70.9%] | 451 [60.3%] | 3,790 [84.5%] | 6,122 [77.6%] | <0.001 |
| Yes | 774 [29.1%] | 296 [39.7%] | 698 [15.5%] | 1,768 [22.4%] |  |
| **Pregnancy loss** |  |  |  |  |  |
| No loss | 2,041 [76.9%] | 572 [76.5%] | 3,269 [72.9%] | 5,882 [74.6%] | 0.002 |
| At least one loss | 614 [23.1%] | 176 [23.5%] | 1,218 [27.1%] | 2,008 [25.4%] |  |
| **Birth past year** |  |  |  |  |  |
| No birth | 2,290 [86.3%] | 641 [85.8%] | 4,177 [93.1%] | 7,108 [90.1%] | <0.001 |
| At least one birth | 365 [13.7%] | 106 [14.2%] | 310 [6.9%] | 782 [9.9%] |  |
| **Respondent's occupation [grouped]** |  |  |  |  |  |
| Not working | 1,539 [58.0%] | 458 [61.2%] | 2,699 [60.2%] | 4,696 [59.5%] | <0.001 |
| Agriculture | 716 [27.0%] | 197 [26.3%] | 1,020 [22.7%] | 1,933 [24.5%] |  |
| Non agriculture | 399 [15.0%] | 93 [12.5%] | 768 [17.1%] | 1,261 [16.0%] |  |
| **Husband/partner's education level** |  |  |  |  |  |
| No education | 675 [25.4%] | 236 [31.6%] | 940 [20.9%] | 1,851 [23.5%] | <0.001 |
| Primary | 864 [32.6%] | 246 [32.9%] | 1,216 [27.1%] | 2,326 [29.5%] |  |
|  | Normal | Underweight | Overweight_Obese | Total | P-value |
| N | 2,655 [33.6%] | 747 [9.5%] | 4,487 [56.9%] | 7,889 [100.0%] |  |
| Secondary | 755 [28.4%] | 203 [27.2%] | 1,544 [34.4%] | 2,502 [31.7%] |  |
| Higher | 361 [13.6%] | 62 [8.2%] | 788 [17.6%] | 1,211 [15.3%] |  |
| Respondent's age at first birth |  |  |  |  |  |
| < 20 | 1,800 [67.8%] | 526 [70.4%] | 2,921 [65.1%] | 5,247 [66.5%] | 0.012 |
| > 20 | 855 [32.2%] | 222 [29.6%] | 1,566 [34.9%] | 2,643 [33.5%] |  |
| Decision-Making Autonomy |  |  |  |  |  |
| Low | 596 [22.4%] | 195 [26.0%] | 747 [16.6%] | 1,537 [19.5%] | <0.001 |
| Mild | 658 [24.8%] | 184 [24.6%] | 1,221 [27.2%] | 2,063 [26.1%] |  |
| Full | 1,401 [52.8%] | 369 [49.4%] | 2,520 [56.2%] | 4,290 [54.4%] |  |

### Test of independence between nutritional Status and explanatory variables of the respondents

The study conducted a bivariate analysis using chi-square tests to assess the association between respondents’ BMI status and the selected explanatory variables. As an initial step in model development, these tests were performed to identify variables significantly associated with the dependent variable. The results of these associations are presented in Table 3. The Table 3 depicts that the socio-demographic characteristics such as age group [chi sq: 404.6301 Pr = 0.000], wealth [541.7548 Pr = 0.000], household size [25.1474 Pr = 0.000] and Urban/rural [25.1474 Pr = 0.000] were all found to be statistically highly significant with non-pregnant married women aged 15-49 in Bangladesh. The respondents [chi sq: 89.2185 Pr = 0.000] and the husband’s highest level of education [161.7438 Pr = 0.000] were also found to be significantly associated with the BMI status of women.

**Table 3.**
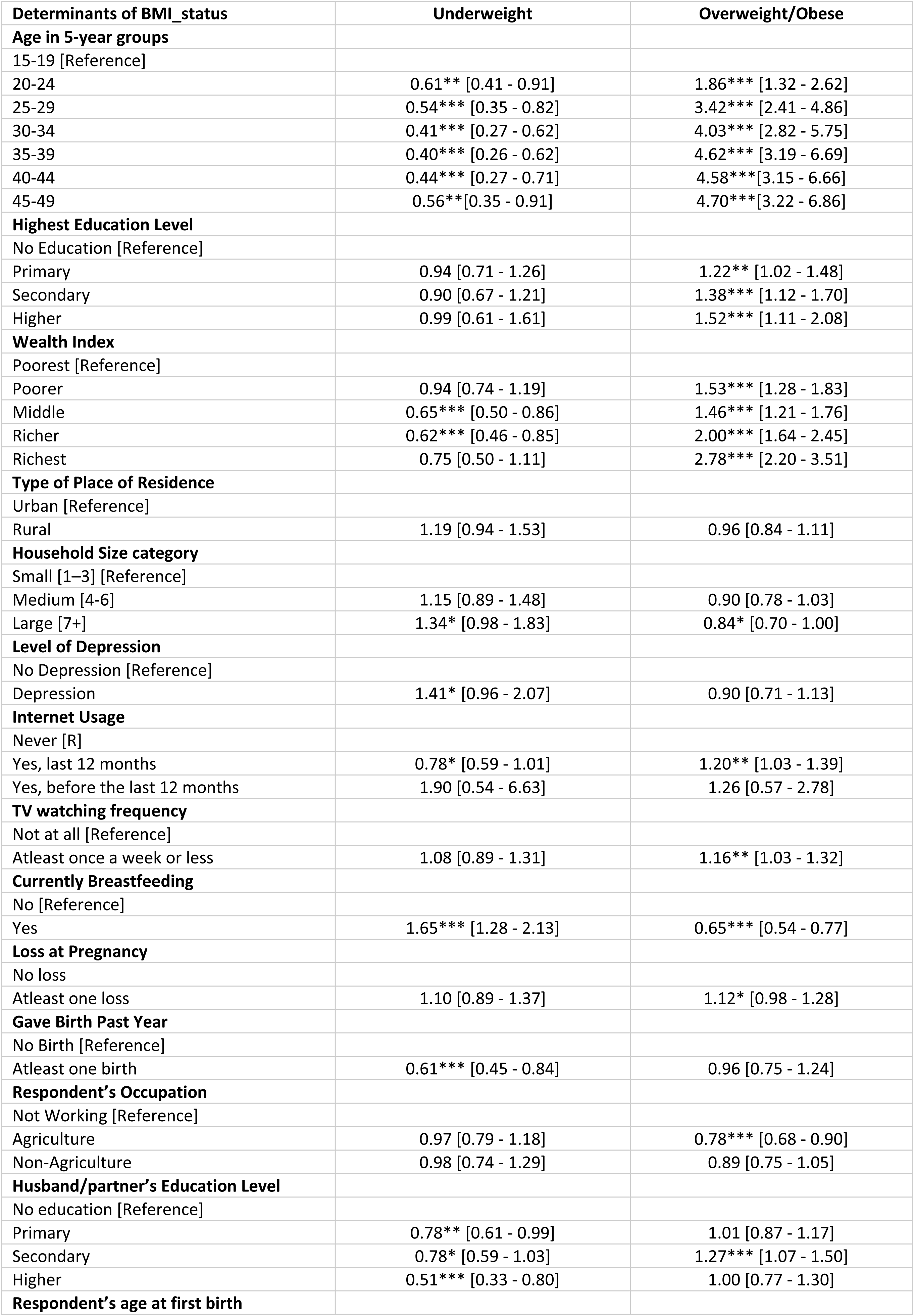
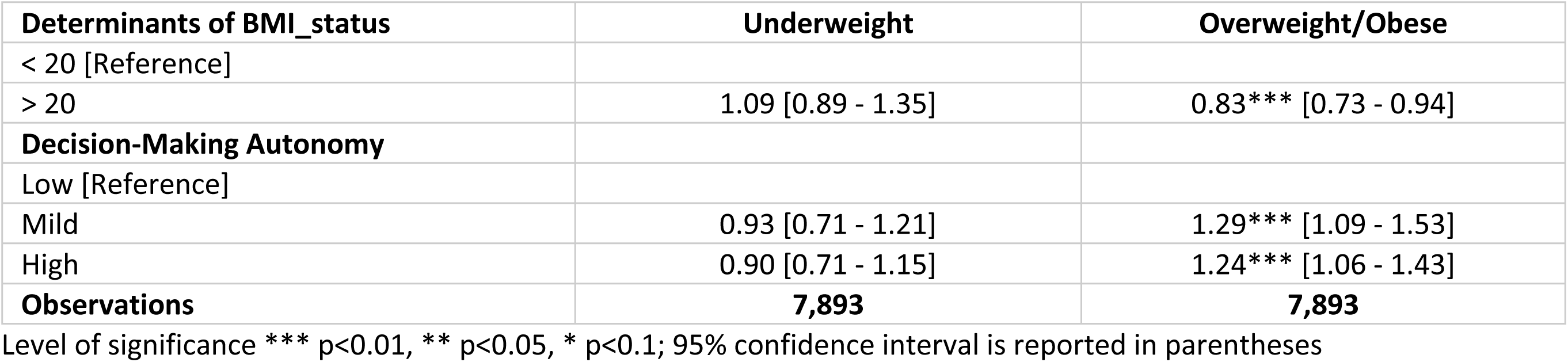
Adjusted odds ratio from the analysis.

Table 3 also shows that reproductive factors such as women’s breastfeeding status, birth past year, age at first birth were significantly associated with the nutritional status of women. Interestingly, this chi-square test of association revealed anxiety level to be insignificant but depression level, women agency - among others- were significantly associated with the BMI status of married women aged 15-49.

### Multivariate multinomial regression to estimate the magnitude of the effects of the explanatory variables on the nutritional status

Variables found to be significantly associated with BMI category in the bivariate analysis were subsequently included in a multivariate multinomial logistic regression model, with normal weight as the reference category, to estimate the magnitude and direction of these associations while adjusting for potential confounders. The results are presented separately for the determinants of underweight and of overweight and obesity to highlight the distinct patterns associated with each outcome.

#### Determinants of Underweight

As illustrated in Table #, sociodemographic characteristics such as age, wealth, household size, respondent’s education and husband’s education significantly explained variations in BMI status. Reproductive factors, including giving birth in the past year, breastfeeding, and age at first birth, also contributed to these differences. The novelty of this study lies in its inclusion of mental health variables, revealing that depression has a strong association with BMI status. In addition, behavioral factors such as internet use and TV watching were found to be significant. Compared to adolescents aged 15–19 years, all older age groups showed significantly lower odds of being underweight. The protective effect was strongest among women aged 30–39 years, who had approximately 60% lower odds of being underweight [aOR for 30–34: 0.40, 95% CI: 0.26–0.61; aOR for 35–39: 0.44, 95% CI: 0.27–0.71]. Marginal effects indicated that women aged 20–24 had a 10.4 percentage point lower probability of being underweight, with this negative association between age and underweight status becoming progressively stronger in the older age groups.

Similarly, women from middle- and higher-wealth households were significantly less likely to be underweight compared to those from the poorest households. Women in the middle and richer wealth quintiles had 35% and 38% lower odds of being underweight, respectively [middle: aOR = 0.65, 95% CI: 0.50–0.85; richer: aOR = 0.62, 95% CI: 0.46–0.85]. Marginal effects indicated that women in the middle wealth group were 5.6 percentage points less likely to be underweight, with the negative association strengthening as wealth increased, reaching 7.2 percentage points lower probability among the richest households.

Household size was an important factor in the nutritional status of married Bangladeshi women aged 15–49. Women living in large households with seven or more members were 3.1 percentage points more likely to be underweight compared to those in smaller households [large households: aOR = 1.34, 95% CI: 0.98–1.83]. Husband’s education also showed a strong association: women whose husbands had education higher than the secondary level had 49% lower odds of being underweight [higher education: aOR = 0.51, 95% CI: 0.33–0.80]. Marginal effects indicated that women were 5.1 percentage points less likely to be underweight if their husbands had completed higher education compared to those whose husbands had no formal education, with the probability decreasing progressively as the husband’s education level increased.

Physiological and health-related factors also influenced women’s nutritional status. Women who had given birth in the past year were significantly less likely to be underweight, with recent childbirth showing 39% lower odds [aOR = 0.61, 95% CI: 0.44–0.84] and reducing the probability by 3.1 percentage points. In contrast, women who were currently breastfeeding were 6.7% percentage points more likely to be underweight or 65% higher odds of being underweight [aOR = 1.65 95% CI: 1.28-2.13].

Women using internet had 22% lower odds of being underweight compared to those who didn’t. [aOR = 0.78 95% CI: 0.59-1.01]. Depression increased the likelihood of being underweight by 3.6 percentage points or in other words, there was 41% higher odds of a depressed woman being overweight/obese. [aOR = 1.41 95% CI: 0.96-2.07].

#### Determinants of Overweight and Obesity

Determinants of overweight or obesity differed notably from those of underweight, with age emerging as one of the strongest predictors. The odds of being overweight or obese increased sharply with age, reaching 4.6-fold higher odds for women aged 35–39 compared to adolescents aged 15–19 [aOR = 4.63, 95% CI: 3.19–6.71]. Marginal effects confirmed this pattern, showing that women aged 35–39 were 37 percentage points more likely to be overweight or obese than those aged 15–19, with the probability increasing at an accelerating rate across successive age groups.

Similarly, respondents’ education level and wealth status showed strong positive associations with overweight/obesity. Women in the richest wealth quintile had nearly threefold higher odds of being overweight or obese compared to those in the poorest quintile [aOR = 2.82, 95% CI: 2.23–3.56], with marginal effects showing 24% increase in probability. Educational attainment also played a significant role, as women with higher education had 53% higher odds of being overweight or obese [aOR = 1.52, 95% CI: 1.11–2.08], corresponding to a 9.1% rise probability compared to women with no education. Similarly, women who had completed higher education were 9.1 percentage points more likely, and those from the richest households were 24.0 percentage points more likely to be overweight or obese than their counterparts with no education and in the poorest households.

Decision-making autonomy was also positively associated with overweight or obesity among married Bangladeshi women aged 15–49. Compared to women with no decision-making agency, those with full autonomy were 5.0 percentage points more likely to be or 24% higher odds of being overweight or obese [aOR = 1.24 95% CI: 1.06 – 1.43]. Several factors were found to reduce the likelihood of being overweight or obesity. Women living in larger households [seven or more members] had a 5.0 percentage point lower probability of being overweight or obese compared to those in smaller households [aOR = 0.84 95% CI: 0.70 – 1.00]. Employment in the agricultural sector was also protective, with women in agriculture having 22% lower odds of being overweight or obese [aOR = 0.78, 95% CI: 0.68–0.90] and a 5.1 percentage point lower probability. Breastfeeding women were 12.1 percentage points less likely to be, or 35% lower odds of being, overweight or obese, reflecting the high energy demands of lactation [aOR = 0.65 95% CI: 0.54 – 0.77].

In contrast, access to digital media increased the likelihood of excess weight gain. Women who used the internet had a 4.9 percentage point higher probability of being overweight or obese [aOR = 1.20 95% CI: 1.03 – 1.39], while those who watched television had a 2.9 percentage point higher probability, compared to non-users [aOR = 1.16 95% CI: 1.03 – 1.32]. Mental health also showed a modest association: women with depression were 4.0 percentage points less likely to be overweight or obese than those without depression, although this result was only significant at the 10% level.

## Discussion

This study found that the weighted prevalence of normal weight, underweight, and overweight or obesity among married women aged 15–49 was 33.7% [95% CI: 32.5%–34.9%], 9.5% [95% CI: 8.8%–10.2%], and 56.9% [95% CI: 55.5%–58.2%], respectively. These findings indicate that overweight and obesity have become highly prevalent, far exceeding the prevalence of underweight, a trend that had been observed in previous studies as well. [Islam, Hossain, Khan, & Rahman, 2021], [Biswas, Uddin,

Mamun, Pervin, & Garnett, 2017]. The 56.9% overweight/obesity rate observed in this study is considerably higher than the ∼25% reported in 2014, reflecting a continuing upsurge in overweight or obesity [Rahman, Rahman, Rahman, & Jesmin, 2019]. Moreover, the 2017–18 BDHS reported that roughly 49% of women are overweight or obese and 12% are underweight. [Rana & Khan, 2022]. The present study’s findings confirm that this trend has continued, with overweight and obesity now comprises most of the burden. Similar patterns were observed in other South Asian countries for instance, India reported 41.2% overweight or obesity [Verma et al., 2023], while Pakistan reported 8.0% underweight and 51.2% overweight or obesity [Waghmare, Chauhan, & Sharma, 2022]. However, these findings suggest that Bangladesh faces a comparatively more severe double burden of malnutrition.

This study revealed that being underweight was more common in women who were younger[adolescents], from poorer and large households, less exposed to media/technology, suffering from depression and more likely to be currently breastfeeding. In contrast, overweight and obese women tended to be older [in their 30s and 40s], more educated, living in wealthier and smaller households, and engaged in more sedentary behaviors [e.g. regular TV/internet use]. These results are consistent with the findings of [Rana & Khan, 2022] [Rahman, Rahman, Rahman, & Jesmin, 2019]

This study identified several demographic and socioeconomic factors that cause undernutrition and overnutrition in women, much in line with patterns reported in the literature. A clear pattern was noticed that as age increases the probability of being overweight and obese increases and that of being underweight declines, a trend supported by many other studies previously [Tanwi, Chakrabarty, Hasanuzzaman, Saltmarsh, & Winn, 2019] [Kamal, Hassan, & Alam, 2015]. Among the age group 15-19 years, it was found that around 11.3% of women were underweight, but only 1.6% were overweight or obese. In contrast, the prevalence of overweight or obese women in higher age groups were mostly between ∼10% to as high as 21%. These findings are well-aligned with established demographic, biological, and socioeconomic explanations. As women age, Basal Metabolic Rate [BMR] declines which can contribute to weight gain [Shimokata & Kuzuya, 1993]. Besides, sedentary behavior -watching TV and using internet-tends to increase with age which further aggravates the obesity problem [McGowan, Powell, & French, 2020].

An interesting observation of this study is that middle-aged women, especially those in the 30–34 and 35–39 age groups, had the highest prevalence of overweight or obesity, at 20.5% and 21.6% respectively. This can be attributed to simultaneous effects of reproductive and physiological aging during midlife, which predispose women to weight gain. [Grammatikopoulou, Nigdelis, & Goulis, 2022]. It was also observed that more educated women in higher wealth quintile had higher odds of being overweight or obese. With each step up in wealth quintile, the probability of a woman being overweight increased dramatically, whereas underweight became correspondingly less common, an observation widely documented in Bangladesh [Gupta et al., 2023] [Khanam et al., 2021] [Biswas, Garnett, Pervin, & Rawal, 2017], similar lower and middle-income countries [Neuman, Finlay, Smith, & Subramanian, 2011] [Bishwajit, 2017] and many sub-saharan African countries. [Neupane, Prakash, & Doku, 2015] [Tebekaw, Teller, & Colón-Ramos, 2014].

In the total sample of this study, 32.4% of the women who were underweight belonged to the poorest wealth quintile, yet only 12.2% of the women who were overweight or obese belonged to the same group. An opposite pattern was observed in the richest wealth quintile where 26.5% were overweight or obese but only 9.4% were underweight. A plausible explanation for this observed association could be women from wealthy households engaging in less physical activity, access to domestic help and having a less healthy dietary habit compared to women in poor households. [Fernald, 2007] [McLaren, 2007].

Education level of women was found to be a significant determinant of overweight or obesity in this study. Many other studies also reported this finding using BDHS data from previous years. [Dey, Zahangir, Faruk, Hossain, & Hossain, 2024] [Khanam et al., 2021] [Sarma et al., 2016].

More educated women earn better and have higher purchasing power of food than women who have had no schooling. Their mode of employment is mostly sedentary which makes them vulnerable to becoming overweight or obese. [Abrha, Shiferaw, & Ahmed, 2016] [Ziraba, Fotso, & Ochako, 2009] [Eriksen, Rosthøj, Burr, & Holtermann, 2015]. A study even showed that Bangladeshi women have the lowest level of physical inactivity when compared to other South Asian women. [Joshi et al., 2007] However, an opposite phenomenon had been observed in developed nations like China and a few OECD countries where higher level of education appeared to be associated with lower likelihood of obesity in women. [Sassi, Devaux, Church, Cecchini, & Borgonovi, 2009] [Aitsi-Selmi, Chen, Shipley, & Marmot, 2013] Another study in conducted in the north-west of Iran reported the same finding [Dastgiri, Mahdavi, Tutunchi, & Faramarzi, 2006]. This was attributed to the fact that educated women might be more knowledgeable about healthy lifestyle and balanced diet intake.

Unlike previous studies, [Rahman, Rahman, Rahman, & Jesmin, 2019] [Biswas, Rahman, Khanam, Baqui, & Ahmed, 2019] education level was not significantly associated with underweight in the current sample. Likely reason behind this finding can be that our sample consists of ever married women and several studies report that married women have better financial stability and household support which in turn protects them from the risk of becoming underweight. [Ikoona et al., 2023] This reasoning was also validated by the finding of this study itself that the prevalence of overweight in married women was almost six times higher than that of underweight.

The study further revealed that a woman’s odds of being underweight decreased significantly with her husband’s education level. This result was consistent with previous research conducted in Bangladesh [Hossain, Khudri, & Banik, 2021] and Ethiopia [Kassie, Abate, Kassaw, & Aragie, 2020]. Unlike previous studies [Hasan, Khanam, & Shimul, 2020], [Dey, Zahangir, Faruk, Hossain, & Hossain, 2024] husband’s education level did not influence a woman’s odds of being overweight in this sample.

This study observed that in comparison to a small size household with 1-3 members, women in medium and large size households had higher chances of being underweight and lower chances of being overweight or obese. [Hasan, Khanam, & Shimul, 2020] This may be attributed to discrimination against women in intra-familial distribution in Bangladeshi households. [Hassan & Ahmad, 1984] [D’Souza & Tandon, 2018]. Another study in India reports similar findings. [Gopaldas, Saxena, & Gupta, 1983] Women working in agriculture showed lower odds of being overweight and obese compared to women who were not working at the time of the survey. A study including 33 low- and middle-income countries showed that women working in agriculture had consistently lower prevalence of overweight.

[López-Arana, Avendaño, Lenthe, & Burdorf, 2013] The rationale behind this finding maybe that agricultural work in lower- and middle-income countries like Bangladesh is relatively less mechanized and more labor-intensive as compared to high-income countries where the availability of modern machineries ease the hard work of labors. Hence, agricultural work may protect against overweight and obesity in LMICs.

This study found that women who watched TV at least once a week had higher odds of being overweight than women who did not. Many studies in Bangladesh reported positive association between women being overweight or obese and TV watching. [Sarma et al., 2016] [Khanam et al., 2021] [Khan & Kraemer, 2009]. Women watching TV are more likely to be exposed to unhealthy food items marketed on TV and are more likely to consume these items. [Alblas, Mollen, Fransen, & Putte, 2018]. TV watching for prolonged hours can also reduce physical activity and energy expenditure. [Hu, Li, Colditz, Willett, & Manson, 2003] [Tucker & Bagwell, 1991]. Hence, these reasons may increase the risk of being overweight or obese.

It was also found that women who used internet in last one year preceding the survey were at higher risk of becoming overweight or obese than women who never used internet. On the contrary, the odds of being underweight was lower for women who used internet in last one year prior to the survey as compared to those who did not. In 2020, a systematic review of 9 cross-sectional studies found that internet use was positively associated with increased odds of being overweight and obese.

[Aghasi, Matinfar, Golzarand, Salari-Moghaddam, & Ebrahimpour-Koujan, 2019] One of the reasons perhaps is that internet use may cause sedentarism, which refers to decreased energy expenditure or physical inactivity. Sedentarism is known to be one of the causes of obesity. [Matusitz & McCormick, 2012] [Vandelanotte, Sugiyama, Gardiner, & Owen, 2009]. Similar association was found in other DHS studies in Nepal and [Rana, Islam, Oldroyd, Samad, & Islam, 2021] and Nigeria [Adeomi, 2025].

A woman’s reproductive factors like age at first birth and whether she gave birth in the past year were also found to affect her nutritional status in this population. Women who gave birth after 20 years of age were at a lower risk of being overweight or obese compared to those who gave birth before turning 20 years old. This can be explained by the fact that many studies found earlier childbirth as a risk factor for increased BMI or obesity. [Damaso et al., 2023] [Danilack, Brousseau, & Phipps, 2018]. Moreover, women who gave at least one birth in the last year prior to the survey were less likely to be underweight than those who didn’t. This is logical as women experience most of the postpartum weight gain in the first year after giving birth. One study found that approximately 75% of women were heavier one year postpartum compared to their pre-pregnancy weight. [Endres et al., 2014] [Sha et al., 2019].

This study revealed that breastfeeding acted as a protective factor against overweight or obesity. In contrast, it was found to be positively associated with the likelihood of being underweight. Many other studies mimicked this finding. [Horta, Rollins, Dias, Garcez, & Pérez-Escamilla, 2022] [Rahman & Sathi, 2020]. Research suggests that lactating mothers are at an increased risk of being underweight due to the higher energy and nutrient requirements associated with lactation [Sserwanja et al., 2021]. Moreover, breastfeeding can also assist with postpartum weight loss. [Rabi et al., 2021].

To the best of our knowledge, this study stated depression as one of the determinants of malnutrition in women for the first time. It revealed that depressed women were at higher risk of being underweight as compared to women who were not depressed. It also found a negative relationship between depression and being overweight or obese. However, the association with overweight was significant at 10% in this sample. The prevalence of depression in married women was found to be 5%. Interestingly, the National Mental Health Survey of Bangladesh [2018-19] recorded the prevalence of depressive disorders in women aged 18 and above 7.9% or 6.5% which is quite close to the finding of this study.

A study revealed that individuals with depression may experience chronic loss of appetite, which subsequently leads to undernourishment. [Tesfa, Jara, Woyiraw, Bogale, & Asrat, 2022] Another study found that being underweight increases the risk of depression in women. [Jung et al., 2017]. Many studies report a U shaped relationship between depression and Body Mass index [BMI] which showed that higher incidence of depression was found in underweight and obese women compared to normal weight and overweight women [Martin-Rodriguez, Guillén-Grima, Aubá, Martí, & Brugos-Larumbe, 2016] [Wit, Straten, Herten, Penninx, & Cuijpers, 2009] [Noh, Kwon, Park, & Kim, 2015]. Compared to women with no agency or decision-making autonomy, those who had mild or full agency were more likely to be overweight. Findings from different studies in Bangladesh, Ghana and Ethiopia indicate that women’s autonomy in household decision making was significantly associated with higher Dietary Diversity Score [DDS] which increased the consumption of a diversified diet among women and other members of the household as well. [Shourove, Meem, Rahman, & Islam, 2023] [Amugsi, Lartey,

Kimani-Murage, & Mberu, 2016] [Kassie, Fisher, Muricho, & Diiro, 2020] Hence, efforts to improve women’s nutrition could be enhanced by strengthening their decision-making power. However, In LMICs, more autonomy doesn’t always translate into healthier outcomes if the food environment is dominated by cheap, calorie-dense, nutrient-poor options and physical activity is limited. An interesting observation found in the sample of this study is that the prevalence of women who are underweight and overweight or obese with full agency is higher than that of women with mild agency.

This study found no significant association of the risk of being underweight or overweight for a woman and the type of place of residence. This could be explained by the fact that after controlling for other socio-demographic factors like age, wealth, education etc. the place of residence itself might not affect the nutritional status of a woman.

## Strengths and Limitations

This study drew data from BDHS which is a nationally representative dataset. It makes the findings relevant at the national level. Moreover, the use of survey weight due to the complex sampling design BDHS uses, provides robust evidence of the factors linked to underweight and overweight status of women. Since BDHS 2022 is the latest round of survey conducted by the DHS program in Bangladesh, this study provides up-to-date evidence on the determinants of women’s nutritional status. BDHS included a mental health module for the first time ever in its 2022 survey. Utilizing the research gap, this study incorporated the mental health dimension to explore the association between women’s depression level and nutritional status. Furthermore, for precision of analysis, Asian specific cut-off points were used to categorize BMI values.

Several limitations of this study should be noted. Due to the unavailability of data on women’s food habits, dietary intake, frequency of smoking and physical activity, these important determinants of BMI could not be added to the model limiting the scope of the study. Moreover, BMI is a crude index which does not account for variations in fat distribution across individuals and populations. [Naser, Gruber, & Thomson, 2006] Therefore, using BMI as a screening tool for obesity is recommended but it should not be used alone for clinical judgement as it may misclassify an individual’s health risk.

Owing to the cross-sectional nature of the data used in this study, establishing a temporal association between the independent variables and the outcome measures is difficult. There is also a possibility of measurement bias as the study factors were measured based on self-report questionnaires. While the findings of this study reflect short-term associations, the long-term effects may differ, an interesting dichotomy warranting further research. Future research can account for these shortcomings to yield more accurate findings.

## Conclusion

Bangladesh is facing a dual burden of malnutrition, especially among married women of reproductive age. Observing the trend of over a decade, it has been seen that the prevalence of overweight or obesity among these women is increasing and that of underweight is declining. The present study reaffirms this trend through its findings of 56.9% prevalence of overweight or obesity and 9.5% prevalence of underweight among married women of age 15-49. The determinants of under- and over-nutrition found in this study were notably distinct. Underweight was concentrated among disadvantaged groups such as younger women- especially adolescents and those from poorer and larger households. On the contrary, overweight and obesity were more common in older age groups, wealthier and smaller households, and among women with higher education and sedentary lifestyles such as frequent TV viewing and internet use. This study discussed various reproductive factors like age at first birth, birth in the past year, breastfeeding status etc. which significantly affected women’s nutritional status. Mental health factors like depression emerged as a risk factor for undernutrition in women. A Woman’s autonomy in decision making also influenced her health significantly.

Finally, improving women’s mental well-being through integrating mental health screening and support into nutrition initiatives is important to break the cycle between depression and poor nutritional status. In summary, national nutrition policies should be revised to tackle the double burden of malnutrition and to ensure healthier futures for Bangladeshi women. Therefore, integrating interventions that tackle both undernutrition and overnutrition are crucial.

## Data Availability

All files are available here: https://dhsprogram.com/data/dataset/Bangladesh_Standard-DHS_2022.cfm?flag=0

## Acknowledgements

Not Applicable

